# Rapid Motor Fluctuations Reveal Short-Timescale Neurophysiological Biomarkers of Parkinson’s Disease

**DOI:** 10.1101/2020.03.04.20024869

**Authors:** Minkyu Ahn, Shane Lee, Peter M. Lauro, Erin L. Schaeffer, Umer Akbar, Wael F. Asaad

**Affiliations:** Department of Neuroscience, Brown University, Providence, RI, 02912; Robert J. and Nancy D. Carney Institute for Brain Science, Brown University, Providence, RI, 02912; The Warren Alpert Medical School, Brown University, Providence, RI, 02903; Norman Prince Neurosciences Institute, Rhode Island Hospital, Providence, RI, 02903; Department of Neurology, Rhode Island Hospital, Providence, RI, 02903; Department of Neurosurgery, Rhode Island Hospital, Providence, RI, 02903

**Keywords:** Parkinson’s disease, neurophysiology, subthalamic nucleus, deep brain stimulation, biomarker, machine learning, motor behavior, oscillations

## Abstract

Identifying neural activity biomarkers of brain disease is essential to provide objective estimates of disease burden, obtain reliable feedback regarding therapeutic efficacy, and potentially to serve as a source of control for closed-loop neuromodulation. In Parkinson’s Disease (PD), microelectrode recordings (MER) are routinely performed in the basal ganglia to guide electrode implantation for deep brain stimulation (DBS). While pathologically-excessive oscillatory activity has been observed and linked to PD motor dysfunction broadly, the extent to which these signals provide quantitative information about disease expression and fluctuations, particularly at short timescales, is unknown. Furthermore, the degree to which informative signal features are similar or different across patients has not been rigorously investigated. Here, we recorded neural activity from the subthalamic nucleus (STN) of patients with PD undergoing awake DBS surgery while they performed an objective, continuous behavioral assessment. This approach leveraged natural motor performance variations as a basis to identify corresponding neurophysiological biomarkers. Using machine learning techniques, we show it was possible to use neural signals from the STN to decode the level of motor impairment at short timescales (as short as one second). Spectral power across a wide range of frequencies, beyond the classic “β” oscillations, contributed to this decoding. While signals providing significant information about the quality of motor performance were found throughout the STN, the most informative signals tended to arise from locations in or near the dorsolateral, sensorimotor portion. Importantly, the informative patterns of neural oscillations were not fully generalizable across subjects, suggesting a patient-specific approach will be critical for optimal disease tracking and closed-loop neuromodulation.

## INTRODUCTION

Parkinson’s Disease (PD), one of the most prevalent neurodegenerative conditions (Pringsheim et al., 2014), is typified by motor and cognitive dysfunction that occur in the setting of pathologically-increased oscillatory neural activity in the basal ganglia (Brown et al., 2001; Priori et al., 2004; Kühn et al., 2005; Weinberger et al., 2006). Lower frequency oscillations, particularly those in the β (∼12–30 Hz) range, have emerged as potential biomarkers for PD motor dysfunction based primarily upon relatively longer timescale observations of abundant β oscillations in the unmedicated PD state and decreased β power in response to therapy (dopaminergic medications or DBS) (Wingeier et al., 2006; Pogosyan et al., 2010; Neumann et al., 2016; Shreve et al., 2016; Neumann and Kühn, 2017; Tinkhauser et al., 2017b). Critically, the relevance of those observations for the moment-to-moment, ongoing manifestation of symptoms on shorter timescales is less well understood. Much as cold temperature is permissive but not causal of snow, it may be the case that β oscillations reflect conditions favorable for the expression of PD symptoms, while separate or additional features of neural activity modulate the immediate dynamics of motor dysfunction.

PD is characterized by a range of signs and symptoms that may include bradykinesia, rigidity, resting tremor, and postural / gait instability, which are heterogeneously manifested across individuals (Lewis et al., 2005; Selikhova et al., 2009; Thenganatt and Jankovic, 2014; Kalia and Lang, 2015). Disease severity is typically measured using the Unified Parkinson’s Disease Rating Scale (UPDRS). Unfortunately, although well-validated (Martínez-Martín et al., 1994; Ramaker et al., 2002; Siderowf et al., 2002), this scale ultimately relies upon subjective patient and clinician assessments and is not intended to capture fluctuations in motor behavior on the timescale of seconds. More objective, quantitative examination of PD patients’ movements revealed that, within individuals, a large dynamic range of movement speed is preserved but shifted towards slower movements (Ballanger et al., 2006; Baraduc et al., 2013). Remarkably, faster movements that approach normal subject velocities (and that maintain normal movement accuracies) can be elicited (Mazzoni et al., 2007), highlighting the impressive residual motor capacity that persists in this condition. Meanwhile, PD patients are notably impaired in their ability to correct for visible target deviations during ongoing movement (Desmurget et al., 2004). Based upon these observations, we hypothesized that quantifying natural motor variability in the context of a simple, continuous-performance, visual-motor task may allow the identification of neurophysiological biomarkers associated with fluctuating motor states.

Our overall strategy was as follows: 1) Engage PD subjects in a continuous motor performance task to elicit natural motor variability and quantify this variability with an array of metrics at short timescales; 2) Apply a machine learning algorithm to determine weights for each of these metrics that maximally differentiate each patient’s motor performance from that of control subjects performing the same task to generate an objective, patient-specific, scalar measure of motor impairment in each short time interval; 3) Examine oscillatory activity in the subthalamic nucleus (STN) while subjects performed this task to determine the manner and extent to which neural signals capture each patient’s fluctuating motor impairment.

## MATERIALS & METHODS

### Subjects

All patients undergoing routine, awake placement of deep brain stimulating electrodes for intractable, idiopathic Parkinson’s Disease between June 2014 and September 2018 were invited to participate in this study. PD patients were selected and offered the surgery by a multi-disciplinary team based solely upon clinical criteria, and the choice of the target (STN vs. Globus pallidus *internus*) was made according to each patient’s particular circumstance (disease manifestations, cognitive status and goals). In this report, we focused on patients undergoing STN DBS (n = 22). Patients were off all anti-Parkinsonian medications for at least 12 hours in advance of the surgical procedure (mean and standard deviation of UPDRS III scores was 50.95 ± 13.6). Approximately age-matched controls (often patients’ partners) also participated in this study (n = 15); patients were aged 47.5–78.5 years (mean 64.3), and controls were aged 48.3–79.2 years (mean 62.6) at the time of testing (t-test comparing the age distributions: p = 0.56). Controls were required simply to be free of any diagnosed or suspected movement disorder and to have no physical limitation preventing them from seeing the display or manipulating the joystick. There was a strong male-bias in the patient population (20M, 2F) and a female preponderance in the control population 3M, 12F), reflecting weaker overall biases in the prevalence of PD and the clinical utilization of DBS therapy (Rumalla et al., 2018).

Patients and other subjects agreeing to participate in this study signed informed consent, and experimental procedures were undertaken in accordance with an approved Rhode Island Hospital human research protocol (Lifespan IRB protocol #263157) and the Declaration of Helsinki. Data from all patients who enrolled and took part in this study are presented here.

### Behavioral Task

We employed a target-tracking task to estimate the degree of motor dysfunction in a continuous fashion. Specifically, while PD subjects reclined on the operating bed in a “lawn-chair” position, a joystick was positioned within their dominant hand, and a boom-mounted display was positioned within their direct line-of-sight at a distance of ∼2-3 feet. The task was implemented in MonkeyLogic (Asaad and Eskandar, 2008; Asaad et al., 2012) and required subjects to follow a green target circle that moved smoothly around the screen by manipulating the joystick with the goal of keeping the white cursor within the circle (Figure 1A). The target circle followed one of several possible paths (invisible to the subject), with each trial lasting 10-30 seconds. Each session consisted of up to 36 trials (∼13 minutes of tracking data), and subjects performed 1-4 sessions during the operation. Control subjects performed this task in an extra-operative setting.

**Figure 1.**
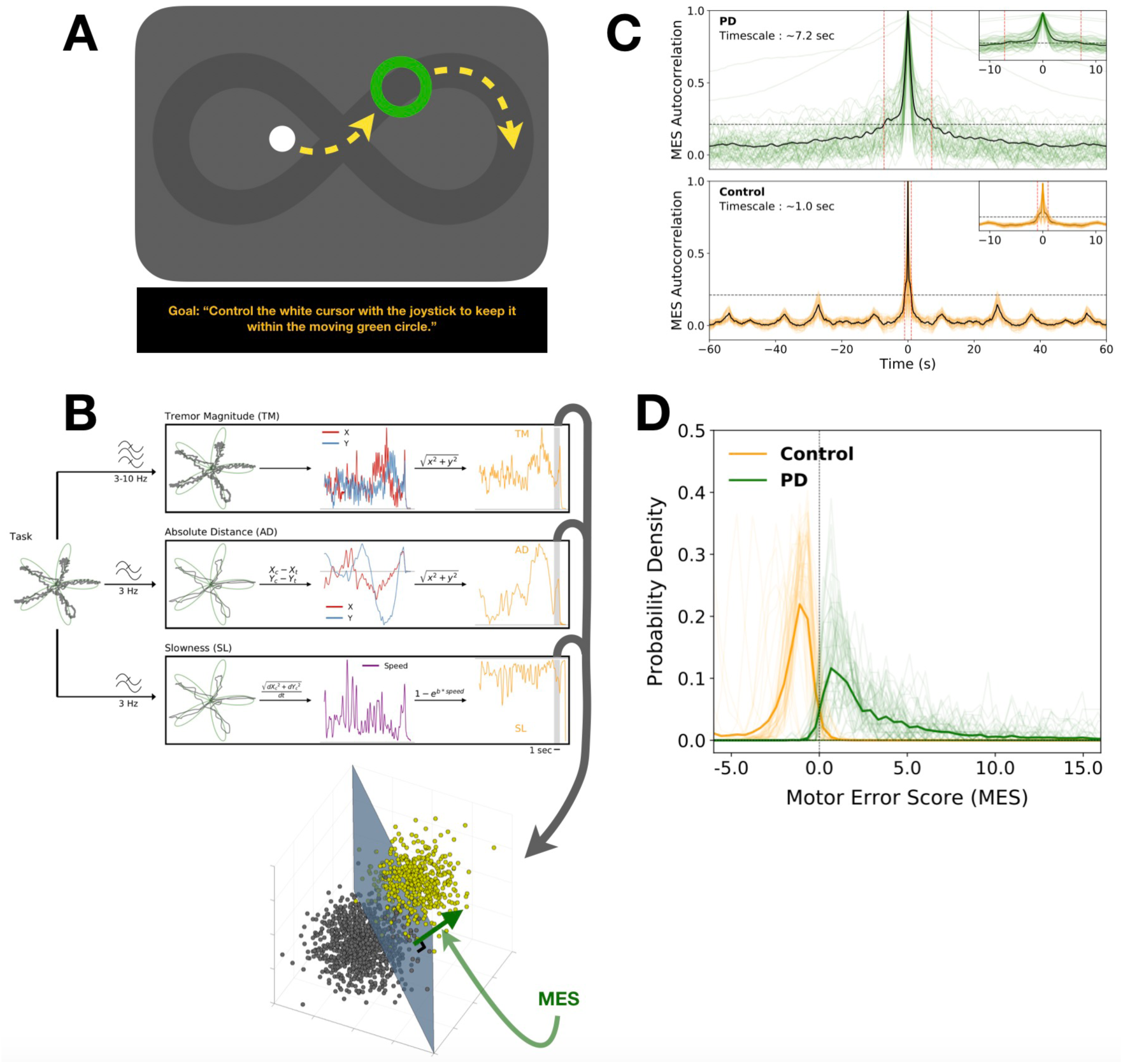
Motor task and behavior. **A**. A continuous motor performance task was used to assess fluctuations in PD motor performance at short timescales. Subjects were instructed to manipulate a joystick to direct a cursor (white) to follow a moving target (green circle) as closely as possible. Yellow arrows depict example trajectories (not displayed to the subjects) with the target following a preset path, (also invisible to the subject, shown here as a dark lemniscate). **B**. A library of 8 motor metrics was created to quantify potential deviations of the cursor from the target. The calculation of three metrics is depicted here (all 8 metrics are defined in Supplementary Figure 1A). The per-epoch estimates of each metric created a distribution in 8-dimensional space; the distribution for a PD subject was compared to those of control subjects via SVM classification. The normal distance from the SVM-defined hyperplane to each point was then defined as the MES for that epoch. **C**. Initially, the MES was calculated in contiguous 100 ms epochs and the autocorrelation was computed to determine the approximate timescale of motor fluctuations. Based on a 3-sigma threshold, the central peak spanned ∼1 second in controls and ∼7.2 seconds in PD subjects. **D**. The PD and control distributions of MES scores at the 7 second timescale were highly discriminable (mean AUC across individuals = 0.99; dark lines = average distributions; lighter lines = individual participants; other timescales shown in Supplementary Figure 1C).

### Motor Metrics & Motor Error Score (MES)

Several measures of motor performance were applied to this task in a small time windows. These measures consisted of tremor (the magnitude of the 3–10 Hz band-pass filtered x- and y-joystick traces) and other metrics calculated after low-pass filtering the x- and y-joystick traces below 3 Hz. The complete library of metrics included tremor magnitude (TM), absolute distance (AD), vector error (VE), slowness (SL), speed difference (SD), excursion difference (ED), vector angle (VA), and correction angle (CA) (Figure 1B and Supplementary Figure 1A). Metrics were initially calculated using 100 ms contiguous, non-overlapping epochs for the behavioral timescale analysis. Metrics were subsequently re-calculated over longer epochs (1–10 seconds) to align with neural data at the relevant timescale for each analysis. Because the motor manifestations of PD are heterogenous, for each PD subject we sought to determine the combination of metric weights that maximally captured their individual motor signs. We applied a linear support vector machine (SVM) algorithm to define the hyperplane in the 8-dimensional metric-space that optimally separated a PD subject’s performance from that of controls. In other words, for each PD subject, the distribution of 8-dimensional data points for each epoch of performance was compared to the aggregate distribution of control subject data points (sub-sampled as needed to maintain equal numbers across groups for SVM binary classification). In all these cases, 100-fold Monte Carlo cross validation was performed using a 2:1 split of the data into training and testing portions. The parameters defining this hyperplane corresponded to the weights applied to the different metrics. The normal distance from each PD subject’s data points (each representing performance during one epoch) to the hyperplane was defined as the “Motor Error Score” (MES).

### Surgical Procedure

Microelectrode recordings (MER) from the region of the STN of awake patients are routinely obtained in order to map the target area and guide DBS electrode implantation. A single dose of short-acting sedative medication (typically propofol) was administered before the start of each procedure, at least 60–90 minutes prior to microelectrode recordings. The initial trajectory was determined on high-resolution (typically 3T) magnetic resonance images (MRI) co-registered with CT images demonstrating previously-implanted skull-anchor fiducial markers. A 3-D printed stereotactic platform (STarFix micro-targeting system, FHC Inc., Bowdoin, ME) was then created such that it could be affixed to these anchors, providing a precise trajectory to each target (Konrad et al., 2011). Microdrives were attached to the platform and then loaded with microelectrodes. Recordings were typically conducted along the anterior, center, and posterior trajectories (with respect to the initial MRI-determined trajectory) separated by 2 mm, corresponding to the axis of highest anatomical uncertainty based upon the limited visualization of the STN on MRI.

MER began about 10–12 mm above the MRI-estimated target, which was chosen to lie near the inferior margin of the STN, about 2/3 of the distance laterally from its medial border. The STN was identified electrophysiologically as a hyperactive region typically first encountered about 3–6 mm above estimated target. At variable intervals, when at least one electrode was judged to be within the STN, electrode movement was paused in order to assess neural activity and determine somatotopic correspondence, as per routine clinical practice. At these times, if patients were willing and able, additional recordings were obtained in conjunction with patient performance of the visual-motor task.

### Neurophysiological Signals & Analysis

Neural signals from the first 4 patients were acquired with FHC tungsten electrodes and an FHC data acquisition system (FHC Inc., Bowdoin, ME, USA), with signals split into a Plexon MAP system (Plexon Inc., Dallas, TX, USA); data for the subsequent 18 patients were recorded using “NeuroProbe” tungsten electrodes and Neuro Omega data acquisition systems (Alpha Omega, Inc., Nazareth, Israel). Electrode impedances were typically 400–700 kΩ.

Patients performed up to 4 sessions of the task, with electrodes positioned at different depths for each session. Electrodes were not independently positionable, so some signals were necessarily acquired outside of the STN. All recorded signals were nevertheless considered and analyzed. Behavioral data from each session were processed through an SVM model, as above, to generate the MES distributions, which were then compared against neural activity acquired simultaneously on individual electrodes.

Data were analyzed in MATLAB (Mathworks, Natick, MA) and Python 3 (python.org). Neural signals from the microelectrodes were initially acquired at 40–44 kHz. Offline, signals were notch filtered at 60 Hz. Time series were z-scored and artifacts above 4 standard deviations were removed. These signals were then downsampled to 1 kHz and bandpass filtered from 3–400 Hz to obtain the micro local field potentials (LFPs). LFPs were bandpass filtered using a fast fourier transform (FFT) convolution with finite impulse response filters. Filters were calculated at 1 Hz intervals with a bandwidth of 2 Hz, covering 3–400 Hz (the use of non-overlapping frequency bins resulted in qualitatively similar results). The power in each band was calculated by multiplying the conjugate of the Hilbert Transform by the filtered signal. We defined six “canonical” frequency bands (θ/α ≝ 4–12 Hz, β ≝ 12–30 Hz, low *γ* ≝ 30–60 Hz, mid *γ* ≝ 60– 100 Hz, high *γ* ≝ 100–200 Hz, “very high frequency” or *vhf* ≝ 200–400 Hz), each consisting of seven sub-bands (Supplementary Figure 2). The width of each band scaled approximately with the frequency because a particular absolute interval in a higher frequency range was likely to be less informative than the same interval in a lower frequency range.

Our goal was to determine which spectral features best corresponded to the MES on short timescales (corresponding to 1– 10 second epochs of task performance and neural data acquisition). To this end, the Pearson correlation of MES with LFP power in particular frequency bands was calculated. Further, to understand if a combination of spectral features might serve as a better marker for MES, continuous decoding of MES was undertaken with a support vector regression (SVR) processing layer using the power-spectral features of the LFP signals. For any particular set of spectral features, SVR estimation accuracy was assessed with 100-fold Monte Carlo cross-validation using a 2:1 training/testing split of each data set. The max-normalized SVR weights were examined to understand the contribution of particular frequency bands to MES classification accuracy (Guyon et al., 2002). Both non-linear (radial basis kernel, *ε* = 0.01) and linear SVR were employed, the former to assess the performance capability of this technique and the latter to allow examination of feature-weight coefficients. Both methods yielded qualitatively similar overall decoding performance across timescales (1–10 seconds).

To make certain machine learning models were not overtrained and/or reliant on spurious associations between behavior and neural activity, a bootstrap procedure was performed in which the correspondence between MES and neural activity in each epoch was randomized and additional SVR models were then generated using the same parameters and cross-validation techniques. Decoding performance of these models was generally quite poor and therefore suggested the performance obtained in the original models was not spurious or due to overfitting. The results of these control analyses are shown in the relevant figures.

SVR coefficients (“weights”) were examined to understand what neural features contributed to MES decoding. Permutation tests were used to determine the significance of these SVR weights and patterns. To assess whether there were clusters of frequency bands with higher or lower weights than expected by chance, a contiguity-sensitive permutation test was performed where the mean SVR weight values were shuffled with respect to frequency over 5000 iterations and the null distribution of weights over at least three contiguous bands was computed. The probability of the observed weight distributions over multiple, adjacent frequency bands could then be determined using this null distribution as reference. To determine if cross-frequency patterns of SVR weight assignments existed, a cross correlation between feature weights for every pair of frequencies was calculated, across SVR models; autocorrelations (points along the diagonal) were excluded from this calculation (diagonal points were filled-in with the means of the adjacent non-diagonal points for the purposes of visualization). Areas of significance within this matrix were computed using a bootstrap procedure in which the assignment of frequency feature weight to LFP signal was shuffled 1000 times to create null distributions for each point on the matrix; the probability of observed values were then calculated with respect to this null distribution.

Unless otherwise specified, statistical tests comparing observed classification or estimation accuracies with those from shuffled MES-neural data sets were performed using a Mann-Whitney U test. Where appropriate, p-values were adjusted for multiple comparisons using the Benjamini-Hochberg procedure (false discovery rate, q = 0.1) (Benjamini, 1995).

### Anatomical Reconstruction of Recording Sites

Patients typically underwent pre-, intra- and post-operative imaging. Preoperatively, stereotactic protocol Magnetic Resonance (MR) images were obtained (Siemens Vario 3.0T scanner) that included T1- and T2-weighted sequences (T1: MPRAGE sequence; TR: 2,530 ms, TE: 2.85 ms, matrix size: 512 × 512, voxels: 0.5 × 0.5 mm^2^ in-plane resolution, 224 sagittal slices, 1 mm slice thickness; T2: SPACE sequence, TR: 3,200 ms, TE: 409 ms, matrix size: 512 × 512, voxels: 0.5 × 0.5 mm^2^ in-plane resolution, 224 sagittal slices, 1 mm slice thickness). Pre-operative, intra-operative (in most cases), and post-operative (in some cases) computed Tomography (CT) scans were also acquired (Extra-Op CT: GE Lightspeed VCT Scanner; Tube voltage: 120 kV, Tube current: 186 mA, data acquisition diameter: 320 mm, reconstruction diameter: 250 mm, matrix size: 512 × 512 voxels, 0.488 × 0.488 mm^2^ in-plane resolution, 267 axial slices, 0.625 mm slice thickness; Intra-Op CT: Mobius Airo scanner, Tube voltage: 120 kV, Tube current: 240 mA, data acquisition diameter: 1,331 mm, reconstruction diameter: 337 mm, matrix size: 512 × 512 voxels, 0.658 × 0.658 mm^2^ in-plane resolution, 182 axial slices, 1 mm slice thickness). MR and CT images were then fused via linear registration using a mutual information algorithm in FHC Waypoint Planner software (version 3.0, FHC Inc., Bowdoin, ME, USA). Localization of the target relied upon a combination of direct and indirect targeting, utilizing the visualized STN as well as standard stereotactic coordinates relative to the anterior and posterior commissures. Appropriate trajectories to the target were then selected to avoid critical structures and to maximize the length of intersection with the STN.

Postoperative MR images (Seimens Aera 1.5T scanner, T1: MPRAGE sequence, TR: 2,300 ms, TE: 4.3 ms, matrix size: 256 × 256 voxels, 1.0 × 1.0 mm^2^ in-plane resolution, 183 axial slices, 1 mm slice thickness, specific absorption rate < 0.1 W/g) were typically obtained 1-2 days after the operation to confirm proper final electrode location.

To reconstruct recording locations, MR and CT images were co-registered using the FHC Waypoint Planner software. The raw DICOM images and the linear transform matrices were exported and applied to reconstructed image volumes using the AFNI command ‘3dAllineate,’ bringing them into a common coordinate space (Cox, 1996). Depths were calculated by combining intraoperative recording depth information with electrode reconstructions obtained from intra- or postoperative images using methods described previously (Lauro et al., 2016; Lauro et al., 2018).

To determine the anatomical distribution of recording sites across patients, pre-operative T1-weighted MR images were registered to a T1-weighted MNI reference volume (MNI152_T1_2009c) using AFNI’s “3dQwarp” command (Fonov et al., 2009). The resulting patient-specific transformation was then applied to recording site coordinates. MNI-warped recording coordinates were then assessed for proximity to the STN as delineated on the MNI PD25 atlas (Xiao et al., 2012; Xiao et al., 2014; Xiao et al., 2017).

### Data Availability

De-identified data and relevant analysis code may be shared upon request for collaborative work with other research groups.

## RESULTS

### Quantification of PD Motor Behavior on Short Timescales

PD patients undergoing awake implantation of STN DBS electrodes (n = 22) and control subjects in the clinic setting (n = 15) performed a tracking task in which they followed an on-screen target using a joystick-controlled cursor (Figure 1A). Because the heterogeneity of motor impairment in PD was unlikely to be associated with some narrow aspect of performance on this task, we defined a library of 8 motor metrics (Supplementary Figure 1A) that tiled the space of potential movement error. We next combined the individual metrics into a single score reflecting the degree of motor impairment within each time window. Specifically, a linear support vector machine (SVM) was applied to the metrics in each epoch to determine, for each patient, a scalar measure of movement quality, the “Motor Error Score” (MES), with higher MES reflecting increased motor impairment (Figure 1B). The MES served as a single composite measure of motor error weighted according to those particular metrics that best distinguished an individual patient’s behavior from that of control subjects.

The PD vs. control MES distributions were distinct (Control: mean ± std = -1.99 ± 2.28; PD: 5.88 ± 10.66, p < 0.0001 by Mann-Whitney U-test). To determine the relevant timescale of motor fluctuations, the MES autocorrelation was computed for PD and control subjects (Figure 1C). The central peak was broader in the PD group, where the timescale incorporating the significant portion (> 3 standard deviations above baseline) was ∼7.2 seconds, while for controls it was only ∼1 second. Importantly, the MES distributions for PD subjects and controls were highly discriminable (Figure 1D shows distributions at the 7-second timescale; other timescales presented in Supplementary Figure 1B); the receiver operating characteristic (ROC) area-under-the-curves (AUC) comparing each PD subject to aggregated control subject performance ranged from 0.95 to 0.99 (mean ROC AUC ± s.e.m. = 0.99 ± 0.01; all p-values < 0.001). These observations that the PD vs. control MES distributions differed in timescale, range, and value support the proposition that the tracking task provided an appropriate platform for detecting PD motor impairment.

### Correlations of STN Neural Activity with Short Timescale Motor Fluctuations

STN local field potentials (LFPs) were recorded from microelectrodes traversing the region of the STN (Figure 2A) from 22 PD patients while they performed the target tracking task. Patients performed 1–4 sessions of the task, with recordings obtained at different depths in each session, for a total of 53 intra-operative sessions. At any particular depth, data were acquired from 2–4 microelectrodes, resulting in a total of 161 recordings from the region of the STN across electrodes, sessions, and patients (Figure 2B). Neural signals from the STN contralateral to the hand used to perform the task were aligned to motor performance (the MES) for analysis (Figure 2C).

**Figure 2.**
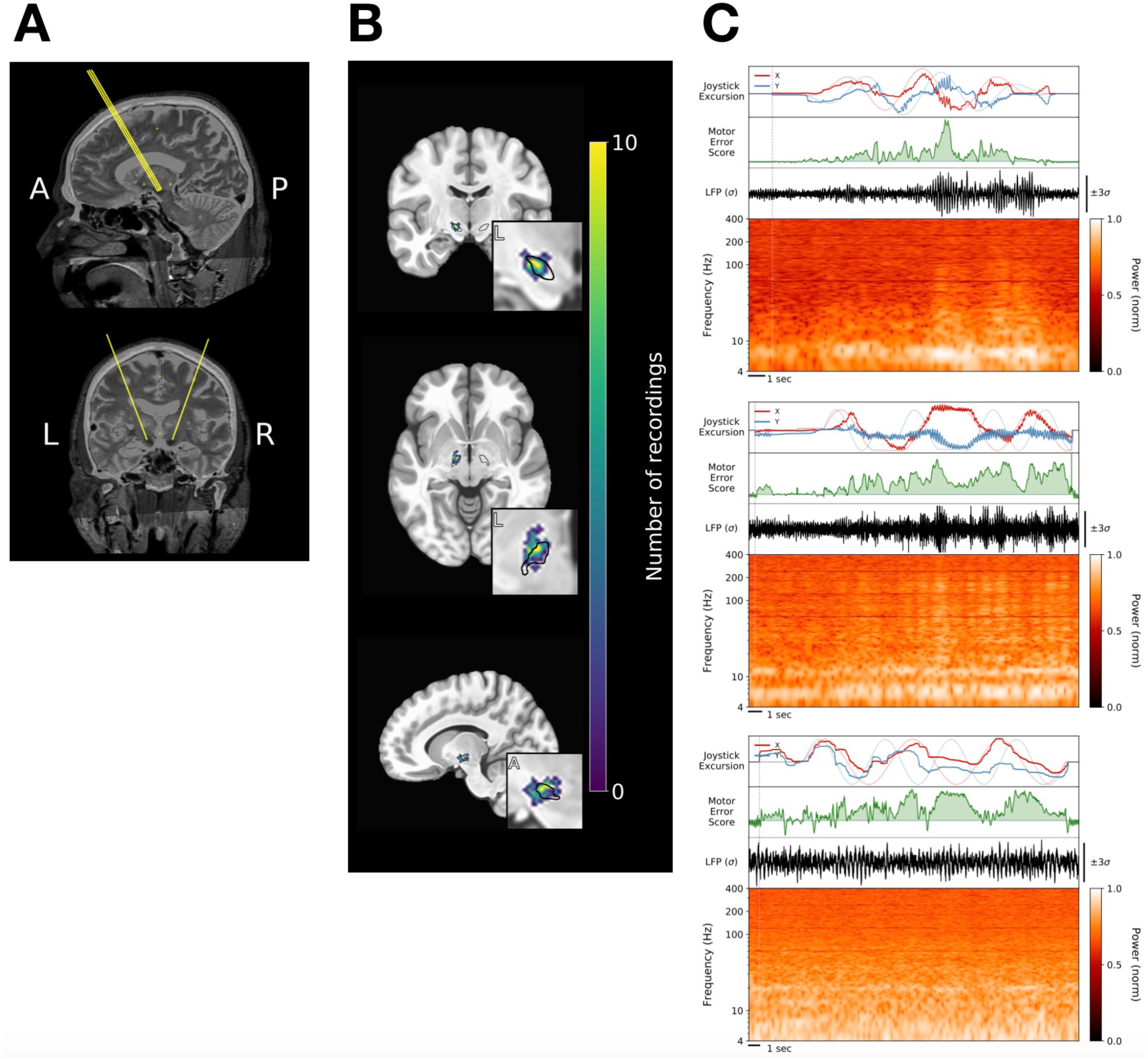
Recording trajectories, locations and signals. **A**. Typical trajectories through the STN. **B**. Density plot of recording locations on an MNI reference volume, with the approximate outline of the STN overlaid in black. Data are plotted for the 20 of 22 patients (140 of 161 recordings) for whom adequate imaging was available. **C**. Three examples of neural data aligned to behavior from three different PD subjects, including decomposition of the LFP signal into the corresponding spectrogram. In the top panel of each example, X- and Y-cursor position (dark red and blue, respectively) are plotted over X- and Y-target position (lighter red and blue, respectively). The MES at corresponding time points is shown below those panels (green). The analytical strategy presented here focused on relationships between spectral features of the STN LFP and the MES.

Oscillations in the β frequency band are considered a likely biomarker for PD symptom expression, and indeed there was often a correlation between β power and motor performance (Figure 3A). However, short timescale correlations between spectral power and MES were not restricted to the β-band. Correlations of MES with θ / α (4–12 Hz) power were generally of similar magnitude to those with β power, and significant correlations were often observed between MES and higher frequency bands as well (Figure 3B). Significant negative correlations were also observed, most often in the *vhf* band (200–400 Hz), suggesting greater power in these higher frequencies may in some cases reflect *better* motor performance. In summary, these correlations revealed information about motor performance was potentially available throughout the tested spectrum (4–400 Hz).

**Figure 3.**
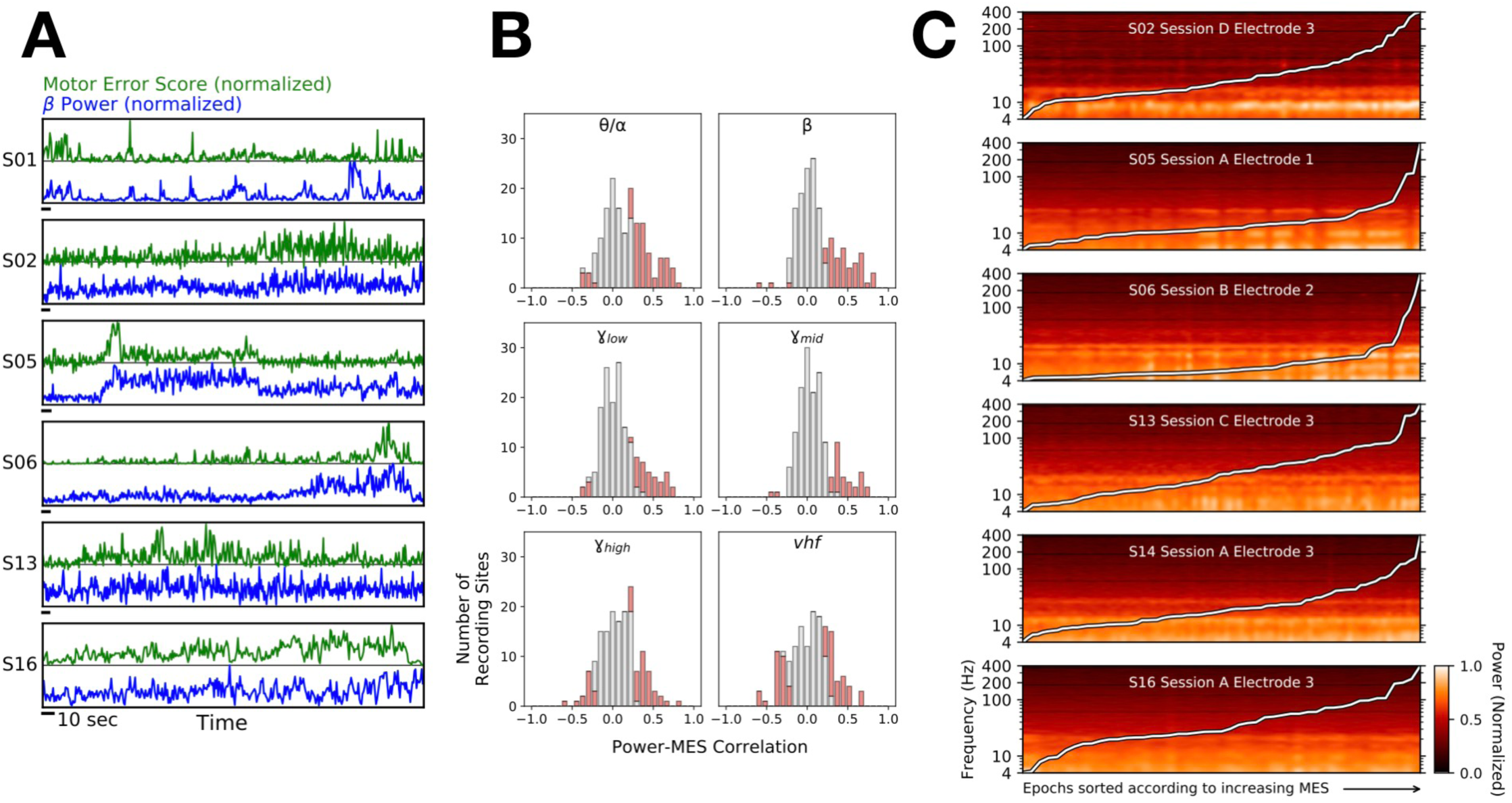
Correlations between STN neural activity and MES. **A**. Examples from six PD subjects demonstrating relationships between β power (blue) and motor performance fluctuations as measured by the MES (green). Horizontal scale bars (black) indicate 10 seconds. **B**. Distribution of Pearson correlation magnitudes between MES and θ/α (4–12 Hz), β (13–30 Hz), low *γ* (30–60 Hz), mid *γ* (60–100 Hz), high *γ* (100–200 Hz) or *vhf* (200–400 Hz) activity across all 22 PD subjects and 161 recordings; red portions of bars denote significant correlations (p < 0.05, corrected for multiple comparisons). Note the increasing presence of signals with significant negative correlations in the higher frequency bands. **C**. Spectrogram examples with epochs (7-second time slices) sorted according to increasing MES. The overlaid white line shows the corresponding MES across epochs.

When the spectrogram was sorted not according to time, but rather according to motor performance, some LFP signals yielded clear patterns where the power in particular frequency bands varied as a function of MES (Figure 3C). There was striking heterogeneity in the particular frequencies that comprised these patterns, but the fact that visually-evident patterns could emerge when spectrograms were sorted in this manner further supported the utility of natural motor fluctuations as a foundation to identify neurophysiological biomarkers of PD behavior.

### Decoding the Quality of Motor Performance using STN LFPs

We next sought to investigate the extent to which STN neural activity could be used to estimate fluctuating motor impairment. Support vector regression (SVR) was applied toward multi-spectral decoding of the MES. Decoding was performed using spectral power across six “canonical” frequency bands (θ/α; β; low *γ*; mid *γ*; high *γ*; *vhf*; Supplementary Figure 2), each divided into 7 sub-bands (yielding a total of 42 spectral features across 4–400 Hz). The data were separated into training and testing portions (2:1) and 100-fold cross-validation was performed. Because the main question related to how well, for any given patient, neural signals might be used to estimate motor impairment, we were interested specifically in the most informative signal from each patient. Note that because recordings were obtained opportunistically when the procedure allowed, and simultaneous electrodes were not independently positionable, many signals were necessarily acquired from suboptimal locations, including outside the STN, so we would not expect all signals to support high-levels of decoding performance.

Indeed, STN neural activity could be used to decode short-timescale motor impairment (Figure 4). Significant decoding capability was evident in every PD subject, with MES decoding accuracy varying from 0.17 to 0.84 (r-values correlating SVR estimates to MES in 7-second epochs: mean = 0.49). Decoding accuracy was generally better at somewhat longer timescales (Figure 5A), reaching a plateau around an 8-second epoch size, but average decoding accuracy with even just 1-second of neural data was nevertheless 0.39 (range: 0.14 to 0.69), increasing to 0.53 (0.29 to 0.88) with 10-second epochs, across all PD subjects.

**Figure 4.**
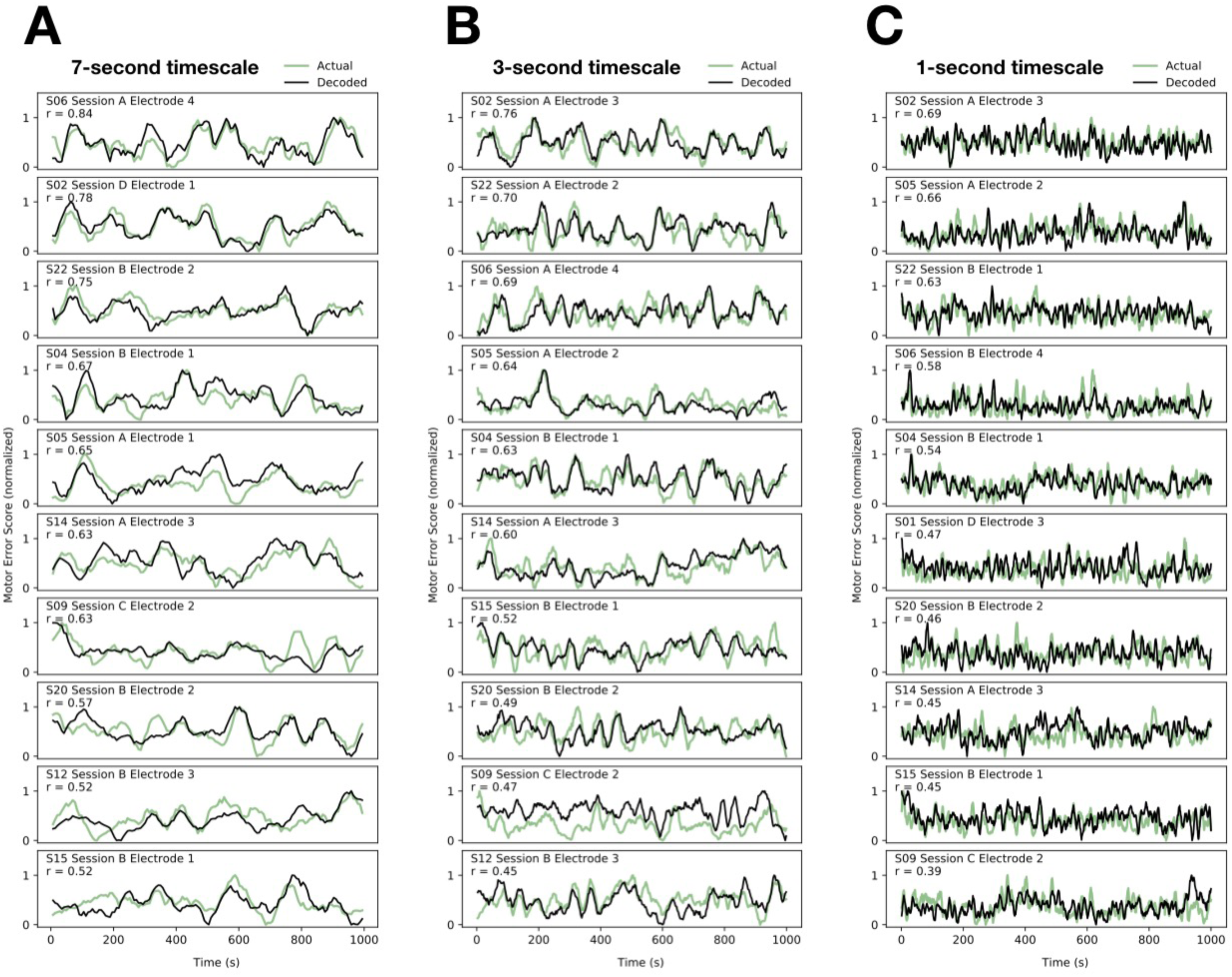
Neural Decoding of Motor Performance: Time-Series. Multi-spectral decoding of motor performance using neural signals obtained from the STN is demonstrated at the 7-second timescale (**A**), the 3-second timescale (**B**) and the 1-second timescale (**C**). In each case, time series for the 10 subjects with the highest decoding accuracies at each timescale are shown (in descending order) to provide a sense of the capabilities of this approach. In each panel, the observed MES (green) is plotted against the SVR-decoded MES (black); SVR estimates are plotted against MES segments with associated neural data that were not used for training the corresponding decoding models.

**Figure 5.**
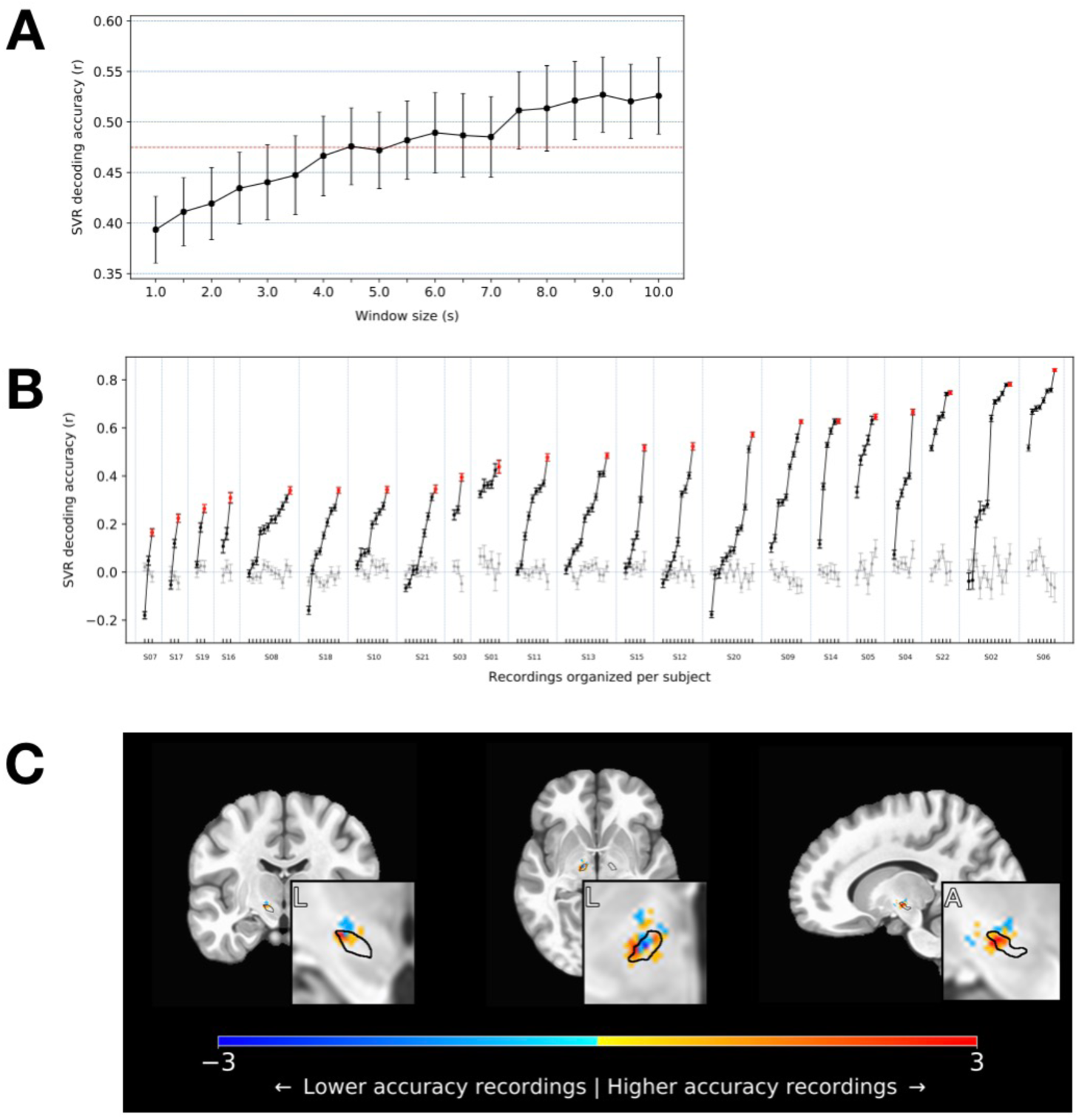
Neural Decoding of Motor Performance: Timescales and Anatomic Localization. **A**. The mean of subjects’ peak decoding accuracies are shown as a function of timescale. Accuracy tended to increase with increasing window size, though a relative plateau in decoding accuracy was observed at about the 8-second timescale. **B**. A wide range of decoder performance was observed across signals, within individuals. Decoding accuracies (r-values, here at the 7-second timescale) are grouped in increasing order by subject. The highest values, reflecting the best achieved decoding performance for each subject, are shown in red. Gray lines show the means of the control analysis in which decoding was performed on neural data shuffled with respect to the MES. **C**. To determine if the best decoding performance tended to arise from signals that were anatomically clustered, recordings yielding the highest (top third) and lowest (bottom third) accuracies were localized on an MNI reference volume (approximate outline of STN overlaid in black), and the differences between these were plotted to reveal the sources of the most informative signals. Coronal, axial, and sagittal slices are shown through the region of peak accuracy, observed near the dorsolateral border of the STN (insets: L = lateral; A = anterior).

There was wide variation in the success of MES decoding across signals even within the same subject (Figure 5B). To determine if this heterogeneity might result from the anatomic source of the neural signals, we next determined the location of the most informative signals by contrasting the top vs. bottom thirds of the decoding models (using all 140 recorded signals across subjects with adequate neuroimaging). The best performing signals arose from the dorsolateral STN, corresponding roughly to the motor subdivision that is classically targeted for DBS therapeutic effect (Figure 5C). Signals recorded within a region-of-interest (ROI) comprising a 2 mm radius around the center of this “hot-spot” (MNI coordinate: x = ±14, y = -12, z = -6) yielded higher decoding accuracies than those recorded outside this ROI (Mann-Whitney U-test: p = 0.001; inside ROI: r = 0.385, n = 28; outside ROI: r = 0.238, n = 112).

In general, broadband, multi-spectral decoding of MES performed better than decoding by individual canonical frequency bands (Figure 6A), consistent with the presence of significant power-MES correlations across all bands, above. To determine the contributions of particular frequency bands to MES decoding, the feature weights of a linear SVR model were examined (Figure 6B). High positive weights (signifying positive correlations between high LFP power and high MES) were typically assigned to the low β band, with less distinct positive peaks visible across the θ/α *and γ* bands.

**Figure 6.**
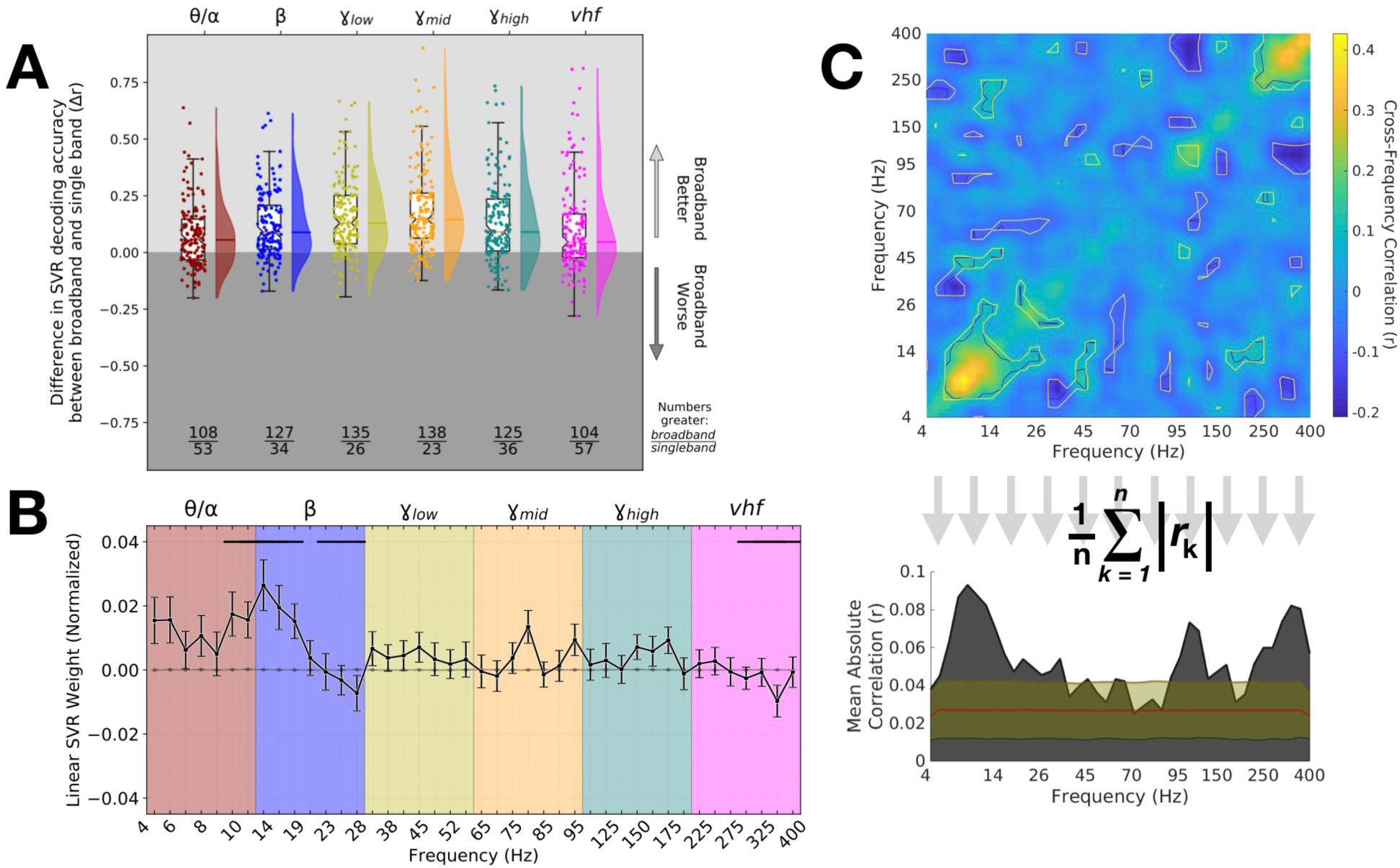
Informative Spectral Features for MES Decoding. **A**. The performance of broadband (4–400 Hz) SVR models to estimate MES is compared against canonical band performance. Each point represents the difference in accuracy for a given LFP signal processed using broadband features vs. features restricted to the indicated band. SVR models were generated and accuracies were determined in identical fashion otherwise. The box plots show the inter-quartile range of each distribution, and the whiskers represent ± 1.5 × the interquartile range. The density of each distribution is depicted in the shaded regions immediately to the right, with the mean value shown by the overlying line. The ratio displayed below each box plot provides the number of recordings for which the broadband accuracy was better (top of ratio) or worse (bottom). The means of all distributions were significantly greater than zero (p < 0.001 by Wilcoxon Signed Rank Test), reflecting generally better performance of the broadband classifier in every case. **B**. The average performance-weighted coefficients of a linear SVR computed for every signal are shown. Positive or negative weights reflected positive or negative relationships with MES. Error bars show the standard errors of the means across subjects, over 100 cross-validation folds. The gray line near the origin shows the mean weights (± standard errors) for the bootstrap SVR feature weight distributions calculated on shuffled MES ↔ neural data. Horizontal bars near the top of the graph represent regions where weights were either greater or lesser than would be expected by chance, based upon a contiguity-sensitive permutation test (p = 0.0002– 0.0064; see Methods). **C**. To determine if cross-frequency patterns of SVR weight assignments existed, a cross correlation between feature weights for every pair of frequencies was calculated, across SVR models. This is plotted (top) along with the mean absolute correlation for a particular frequency in relation to all others (bottom). The contours on the correlation matrix depict significance (α) levels of 0.001 and 0.01 (dark and light contours, respectively) computed using a bootstrap permutation test to create “null” distributions for each point on the matrix (see Methods). The overlaid region on the lower area plot includes the mean (red line) and mean ±3 standard deviations (yellow line) as determined using the same bootstrap distributions as for the correlation matrix. Several significant peaks are visible above this range, most notably in the θ/α and low β ranges, in the low/mid *γ* range, and in the *vhf* range, confirming cross-frequency interactions were most consistently observed across these ranges.

Meanwhile, negative weights were more typically assigned to the high β band and *vhf* band, suggesting higher LFP power in these frequency ranges was often associated with better motor performance (lower MES).

We hypothesized patterns of weight assignments might exist across frequencies, such that weights assigned to particular bands might co-vary with weight assignments to other bands; the existence of such higher-order patterns would strengthen the notion that those involved bands were consistently meaningful, and might provide insight into potentially shared oscillatory mechanisms. Indeed, significant positive correlations were observed across adjacent bands in the θ/α *and* β ranges, and within the *vhf* band (Figure 6C); this reflected the coexistence of positive weights in the former and negative weights in the latter. In addition, local positive correlations were observed in the mid/high *γ* band as well. Interestingly, relatively strong negative correlations were observed between the vhf and mid/high *γ* bands, implying effective decoding models incorporating low vhf power relied also on increased *γ* power. Likewise, a relatively prominent negative correlation was observed between the low *γ* and θ ranges. A variety of other significant correlations were observed as well, suggesting these potentially interesting cross-frequency interactions may underlie the spectral “fingerprint” of PD motor dysfunction.

However, these averaged patterns of SVR weights belied noteworthy individual variability. A “generic” model constructed from these mean weights faired poorly in estimating the MES (Figure 7A); specifically, the generic SVR decoder significantly outperformed patient-optimized decoders for only 1 of the 22 PD subjects, whereas the patient-optimized decoders significantly outperformed the generic decoder in 15 of 22 cases (by Wilcoxon paired test with α = 0.05, corrected for multiple comparisons; 6 cases demonstrated no significant difference between these techniques).

**Figure 7.**
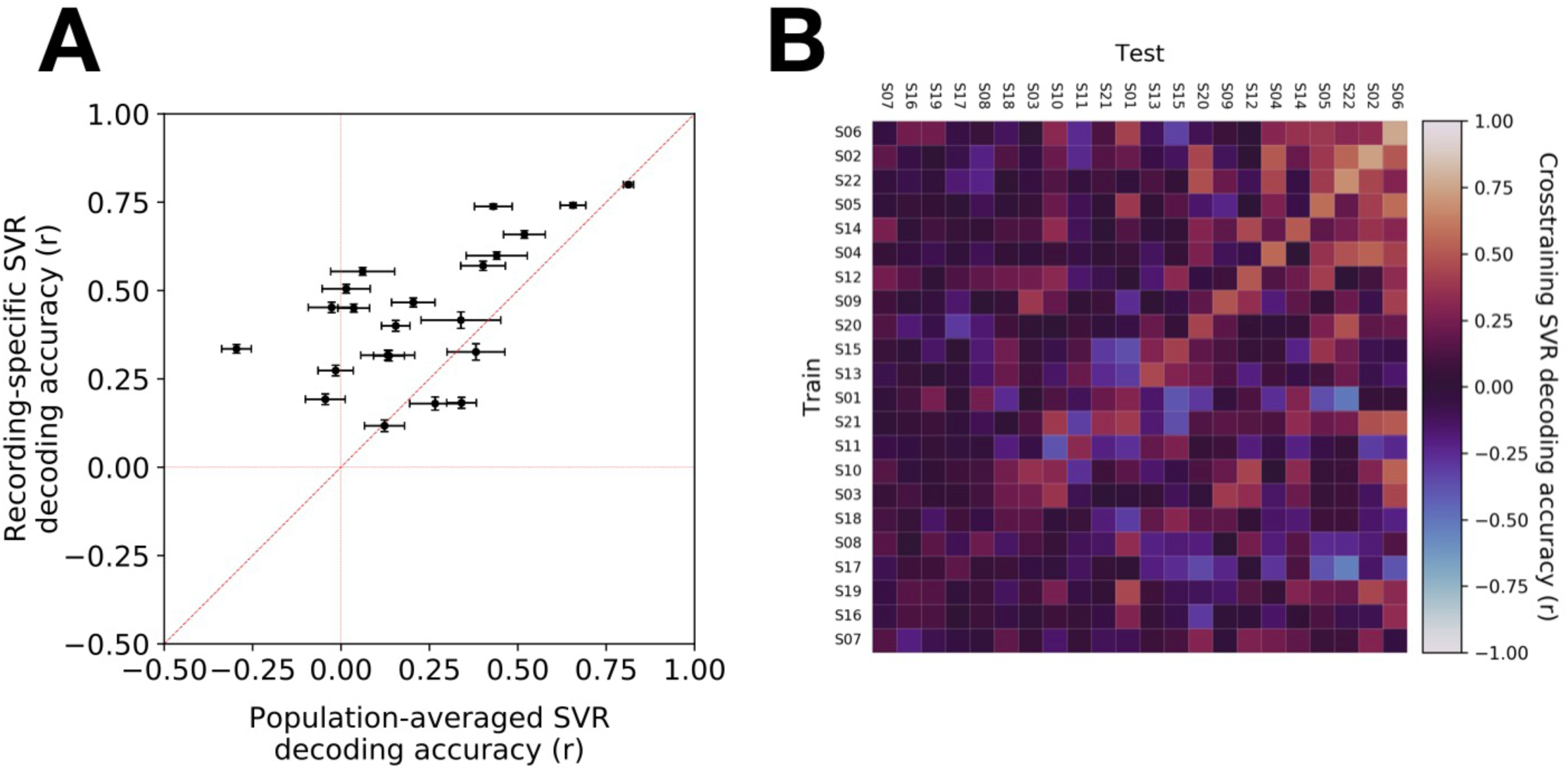
Non-generalizability of MES decoding models. **A**. A “generic” classifier was constructed using the average, performance-weighted linear SVR model coefficients, across patients, for each frequency feature. The performance of the generic decoder (x-axis) was compared against SVR decoders generated for each subject (y-axis). Patient-specific decoders generally performed better than the generic decoder. **B**. To determine if there might be particular decoding models derived from individual subjects that generalized well to others, decoders were trained on one subject’s data and tested on another’s data to estimate the latter’s MES, across all pairs of subjects. The results are plotted in a cross-decoding matrix where the best result across each pair of subjects is shown (selected from among all the recording pairs between those subjects). The cases along the diagonal (where the same subject’s data were used for training and testing) are referred to as “autologous,” whereas the off-diagonal cases (reflecting training and testing across subjects) are termed “homologous” conditions. Because there are more combinations of homologous electrode pairs across subjects than there are autologous cases within subjects, this analysis potentially was biased against the autologous condition. Nonetheless, the best-performing cases were typically found on the diagonal. Furthermore, the lack of horizontal banding in the matrix demonstrates there was no “special” decoder that succeeded in generalizing across patients, and the lack of vertical banding demonstrates there was no particular patient whose neurophysiological “fingerprint” was sufficiently simple that it could be estimated successfully using some broad set of homologous decoders that happened to emphasize some simple feature.

Likewise, patient-specificity was evident in a cross-decoding analysis where the training and testing of SVR models was conducted across pairs of subjects (Figure 7B). For any given patient, decoding models trained on that individual’s signals (“autologous” conditions, on the diagonal) usually outperformed decoders trained using another’s signals (“homologous” conditions, off-diagonal). It may be relevant, however, that the highest-performing quadrant of this matrix (top-right) had somewhat higher cross-decoding success than the lowest-performing quadrant (bottom-left) suggesting that, even though the autologous decoders (along the diagonal) had superior accuracy, there was nevertheless evidence for slightly improved generalizability across the best-performing multi-spectral SVR models and subjects.

## DISCUSSION

We examined endogenous fluctuations in movement quality to elucidate the neural correlates of motor dysfunction at short timescales. We used a naturalistic task that elicited goal-directed action such that deviations from the intended target could be quantified to provide an objective measure of immediate motor impairment in PD subjects. Multi-dimensional assessment of behavioral performance on this task discriminated between PD subjects and controls in a highly accurate manner and organized neural activity in a sufficiently meaningful way that it could be used to decode the quality of movement. Specifically, the power spectral features of the STN LFP correlated with fluctuating behavior, and enabled the quality of motor performance to be decoded by SVR using as little as 1-second of neural data from a single microelectrode. Importantly, decoding relied upon a broad range of frequency bands in a patient-specific manner. The links between neural rhythms and motor dysfunction on these short timescales suggest that such oscillations do not simply set a broad state within which motor performance fluctuates, but rather that the rapid dynamics of these oscillations correlate directly with impairments in immediate, ongoing motor behavior.

With a few notable exceptions, prior work examining the relationships between oscillatory activity and PD manifestations considered only longer timescales, corresponding to on vs. off medication states, or on vs. off DBS stimulation. Some of what those studies found mirrors what we observed here at shorter timescales, such as a possible distinction between high and low frequency β activity (Fogelson et al., 2006; Litvak et al., 2011; Little et al., 2013; van Wijk et al., 2016), the importance of sub-β (i.e., θ/α) activity (Hirschmann et al., 2016), and the possible relationship between higher-frequency activity and more fluid motor output (Sharott et al., 2014; Swann et al., 2016). Here we show that power across a wide gamut of frequencies, including but not limited to these, were significantly associated with PD motor dysfunction.

Furthermore, combining multiple spectral features significantly improved motor performance decoding. Interestingly, we observed novel interactions between activity across different portions of the spectrum, such as negative correlations between α / low-β and high-β, and between mid/high *γ* and *vhf*. Such interactions may provide clues about the underlying mechanisms that generate PD-relevant activity.

The few notable studies that have examined PD neural activity in relation to shorter-timescale motor behavior have focused on specific frequency bands. For example, β “bursts” immediately preceding movement were found to be associated with impaired movement dynamics (Torrecillos et al., 2018) and these bursts were linked to bradykinesia during repetitive movements (Lofredi et al., 2019), consistent with our results in the low, but not high, β range. Meanwhile, *γ* activity was observed to correlate positively with movement velocity at short timescales (Lofredi et al., 2018). In contrast, we found that *vhf* activity was most consistently reflective of effective motor output.

The increased performance of multi-spectral motor performance decoding, compared to single-band decoding, may result from better estimation of multiple dimensions of motor signs, or it could reflect better estimation of individual motor features that have representations in multiple parts of the spectrum. In addition, the wide range of informative frequency bands could reflect cognitive processing related to the recognition, evaluation, or attempted correction of motor errors (Lardeux et al., 2009; Tan et al., 2014). This might help to explain the presence of significant information about motor performance throughout the recorded region, outside the dorsolateral, sensorimotor “hot spot.”

The precise pattern of informative oscillations varied across individuals. Some of this may reflect anatomical differences (inexact correspondence of STN subregions recorded across patients). For example, the distributions of β and α activities may differ topographically across the STN (Horn et al., 2017). Here, neural recordings were performed opportunistically when the operative procedure allowed; this may have been in part responsible for the variability of symptom state decoding accuracy and feature weights across patients. Furthermore, there is likely an interaction between spatial location and symptomatic subtype in determining the particular pattern of neural oscillations (Sharott et al., 2014; Telkes et al., 2018). Nonetheless, we observed relatively poorer generalizability of neural decoders across individuals, suggesting that while there may be some common substrates of particular symptoms within certain regions of the STN, there is nevertheless significant heterogeneity that may be harnessed to yield more optimal decoders using patient-specific biomarkers. Furthermore, because microelectrode-derived LFP recordings detect more spatially-restricted signals, our use of micro-LFP signals rather than macro-electrode (e.g., DBS electrode) LFPs may have highlighted individual differences. Consistent with this, prominent STN signal heterogeneity has been observed using experimental DBS electrodes with smaller electrical contacts (Bour et al., 2015).

Our approach to decoding ongoing motor dysfunction using neural activity may potentially be further optimized. On the front-end, while the continuous visual-motor tracking task succeeded in differentiating the behavior of PD subjects from controls to a high degree, it is unlikely this task captures the full “ground truth” of motor disability, even for the single upper extremity for which it was evaluated. Improved methods to quantify motor signs, including perhaps over the entire span of bodily movement, may further enhance decoding accuracy.

On the back-end, for MES decoding, we restricted the input feature space to LFP power at different frequencies. This ignored significant information that is likely present in the phases of these oscillations, particularly when combined with amplitude information (Lopez-Azcarate et al., 2010; Yang et al., 2014; van Wijk et al., 2016; Telkes et al., 2018). Further optimization of the feature sets and algorithms (e.g., artificial neural network approaches) may yield more precise decoders of motor impairment. Nonetheless, our use of simple features and models for neurophysiologically-based MES decoders allowed us to focus on the links between specific aspects of neural activity and quantified behavior in a relatively straightforward manner.

Short timescale neurophysiological biomarkers of PD motor dysfunction may enable several important, practical applications. Specifically, the ability to estimate and track motor quality continuously over time would allow objective feedback about real-world, “in the wild” disease burden, and so may facilitate adjustments of medical therapy. Likewise, closed-loop DBS, in which stimulation is delivered dynamically, as needed, requires a control signal that reflects the immediate or impending presence of symptoms. Our results suggest that proposed strategies focusing largely on broad β-band activity may be suboptimal and not fully generalizable across patients and STN sites. Note also that STN β decreases just before and during movement (Foffani et al., 2004; Kühn et al., 2004; Yugeta et al., 2013), including tremor (Qasim et al., 2016), so its use as a surrogate for PD motor dysfunction during ongoing behavior might be limited (Johnson et al., 2016). Furthermore, β oscillations are not necessarily pathological, even in PD (Deffains et al., 2018), so stimulating simply to disrupt β might have unanticipated adverse influences on motor behavior. That and other work (Tinkhauser et al., 2017a; Tinkhauser et al., 2017b) have suggested targeting longer β episodes might more selectively address PD symptoms. We show here that patient-specific, multi-spectral decoding of motor impairment can further refine the strategy for implementing closed-loop DBS. Ultimately, a better understanding of the links between neural activity and motor behavior on the relevant timescales, and novel algorithms to fully exploit these links, will guide the design of more advanced therapies to realize the full potential of neurophysiology and neuromodulation for optimal patient benefit.

## ACKNOWLEDGEMENTS

We are grateful for the generous participation of our patients in this study. We thank Kelsea Laubenstein-Parker for technical assistance, David Segar, Tina Sankhla, and Daniel Shiebler for early implementation of a version of the motor task, Karina Bertsch for administrative support, and Ann Duggan-Winkle for clinical support. WFA, MA & SL designed the experiment; MA, SL, PML, JG, ELS, UA and WFA conducted the experiment; MA, SL, PML and WFA performed the analyses; WFA, SL and PML wrote the manuscript; all authors reviewed and edited the manuscript.

## Abbreviations

AUC: Area Under-the-Curve
CT: Computed Tomography
DBS: Deep Brain Stimulation
LFP: Local Field Potential
MER: Microelectrode Recordings
MES: Motor Error Score
MRI: Magnetic Resonance Imaging
PD: Parkinson’s Disease
ROC: Receiver Operating Characteristic
STN: Subthalamic Nucleus
SVM: Support Vector Machine
SVR: Support Vector Regression
UPDRS: Unified Parkinson’s Disease Rating Scale

## LIST OF SUPPLEMENTARY MATERIALS

Supplementary Figure 1. *Motor metrics and their correlations in PD subjects and controls*.

Supplementary Figure 2. *Frequency range definitions*.

Supplementary Video. *Examples of tracking task performance by PD and control subjects*.

## FIGURES & LEGENDS

**Supplementary Figure 1.**
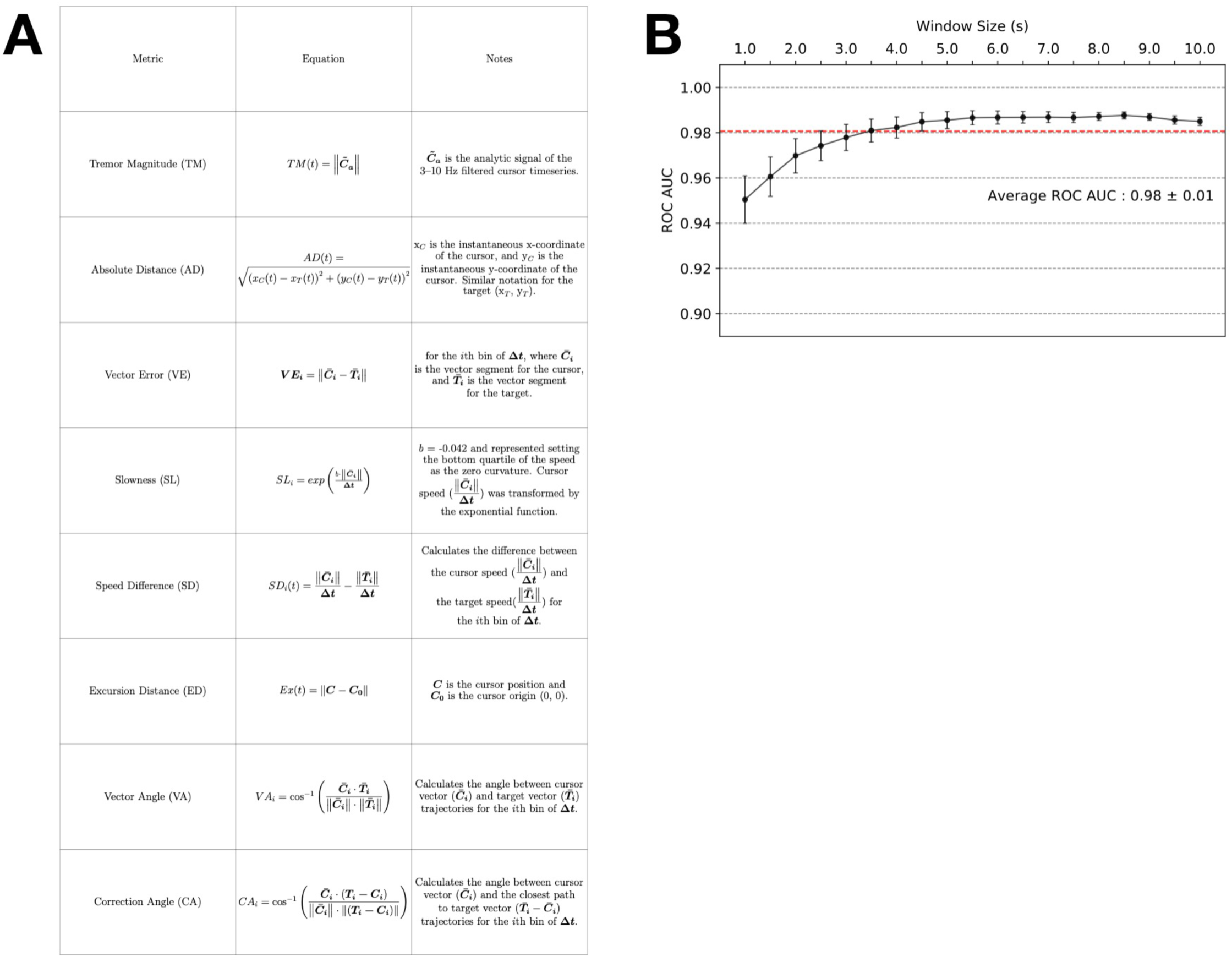
Motor metrics and classification of PD subjects vs. controls across timescales. **A**. The eight motor metrics assessed within each epoch of task performance. **B**. The multi-metric approach to behavioral assessment, yielding the MES, enabled good discrimination between control subject performance and PD subject performance at all tested timescales. Shown are the means ± standard errors for all subjects and sessions at each timescale.

**Supplementary Figure 2.**
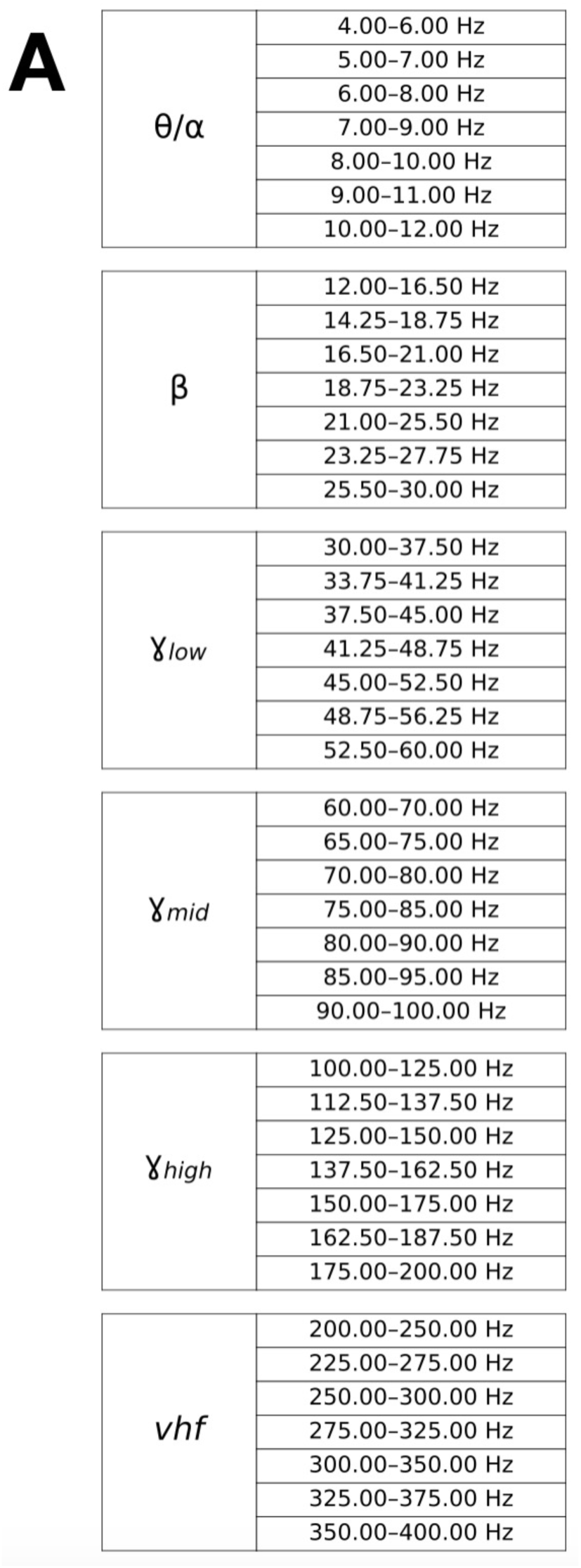
Frequency range definitions. **A**. The 4–400 Hz range was divided into 6 “canonical” bands, which were each then subdivided into 7 sub-bands to serve as finer-grained features for MES decoding using neural activity.

**Supplementary Video**. *Examples of tracking task performance by PD and control subjects*. Shown is one trial of tracking task performance by four subjects (three PD, one control). The top panels show target and cursor trajectories with a disappearing trail (not shown during actual task performance) to visualize performance more clearly. The MES is plotted in the bottom panels.

